# How are adversities during COVID-19 affecting mental health? Differential associations for worries and experiences and implications for policy

**DOI:** 10.1101/2020.05.14.20101717

**Authors:** Liam Wright, Andrew Steptoe, Daisy Fancourt

## Abstract

**Importance:** Multiple data sources suggest that COVID-19 is having adverse effects on mental health. But it is vital to understand what is causing this: *worries* over potential adversities due to the pandemic, or the toll of *experiencing* adverse events.

**Objective:** To explore the time-varying longitudinal relationship between (i) worries about adversity, and (ii) experience of adversity, and both anxiety and depression and test the moderating role of socio-economic position.

**Design:** Longitudinal cohort study

**Setting:** Community study

**Participants:** A well-stratified sample of UK adults recruited into the UCL COVID -19 Social Study (a panel study collecting data weekly during the Covid-19 pandemic) via a combination of convenience and targeted recruitment. The sample was weighted to population proportions of gender, age, ethnicity, education and geographical location.

**Exposures:** Worries or experiences of adversities during the COVID-19 pandemic

**Outcomes:** Anxiety (GAD-7) and depression (PHQ-9)

**Results:** Data were analysed from 41,909 UK adults (weighted data: 51% female, aged 18-99) followed up across 6 weeks (178,430 observations). Using fixed effects regression was used to explore within-person variation over time, cumulative number of worries and experience of adversities were both related to higher levels of anxiety and depression. Number of worries were associated more with anxiety than depression, but number of experiences were equally related to anxiety and depression. Individuals of lower socio-economic position were more negatively affected psychologically by adverse experiences.

**Conclusions & relevance:** Measures over the first few weeks of lockdown in the UK appear to have been insufficient at reassuring people given we are still seeing clear associations with poor mental health both for cumulative worries and also for a range of specific worries relating to finance, access to essentials, personal safety and COVID-19. Interventions are required that both seek to prevent adverse events (e.g. redundancies) and that reassure individuals and support adaptive coping strategies.

**Key points:** *Question:* How do *worries* over potential adversities due to the COVID-19 pandemic, or the toll of *experiencing* adverse events affect mental health?

*Findings:* Cumulative number of worries and experience of adversities were both related to higher levels of anxiety and depression during COVID-19, especially amongst individuals of lower socio-economic position.

*Meaning:* During a pandemic, interventions are required that both seek to prevent adverse events (e.g. redundancies) and that reassure individuals and support adaptive coping strategies.

## Introduction

The global pandemic of COVID-19 is leading to increasing experience of adversities, from infection and serious illness due to the virus, to financial shocks such as loss of employment and income, to challenges in accessing food, medication or accommodation, to adverse domestic experiences such as abuse ^1,2^. These experiences echo those reported during previous epidemics ^3^. However, their effects are causing even greater concern than in epidemics previously due to the global spread of the virus, the scale of lockdown measures that are proving necessary to contain the spread (which are having major effects on economies), and the long time-scale being projected for the pandemic^1,4^.

In particular, there are concerns that COVID-19 will have substantial and lasting effects on mental health ^5^. Already, reports are emerging of a parallel epidemic of fear, anxiety and depression ^6^. But at present, it remains unclear what is triggering these adverse psychological effects: *worries* over potential adversities due to the virus, or the toll of actually *experiencing* adverse events. Literature suggests that experiencing adversities such as ill-health, financial problems, and challenges meeting basic needs is associated with poor psychological outcomes including anxiety, depression, post-traumatic stress, and broader distress ^7,8^. This has been found to apply to situations in epidemics too ^9^. However, it is not just experiencing adversities that can have such effects; even worries about experiencing adverse effects can negatively impact on mental health. For example, experiencing daily worries is associated with depressive symptoms both in the short-term and over several years ^10,11^. This has been shown for a range of worries, including those relating to health and finances ^12,13^. In fact, worries and other negative reactions to an event have in some instances been found to be more important in predicting mental health and wellbeing than experiencing the event itself ^14^. It is vital to ascertain whether it is worries of adversity or experiences of adversity that are most strongly linked to declines in mental health as each require different types of support or interventions to prevent or mitigate their effects. For example, if worries are most strongly associated with poor mental health, then provision of greater public reassurance or individual interventions such as online cognitive behavioural therapy programmes could be made more available to people. In contrast, if experience of adversity shows greater associations with poor mental health, then interventions that provide more tangible and material support (such as further financial relief measures) may be key.

Additionally, there are worries that adversities will exacerbate existing inequalities within societies by disproportionally affecting individuals of lower socio-economic position (SEP) ^1,2^. These individuals are more likely to experience adverse events during the pandemic, as well as more likely to have poorer mental health in the first place ^3,15^. Low SEP individuals may also have fewer material and psychosocial resources available to deal with adversity ^16^, and studies specifically looking at the effect of adversity on mental health have shown that there is socio-economic variation in the consequences of adversity ^17^.

Therefore this study used a large, longitudinal dataset of weekly experiences during the early weeks of the lockdown due to COVID-19 in the UK to explore the time-varying longitudinal relationship between (i) worries about adversity, and (ii) experience of adversity, and both anxiety and depression. Further, it sought to ascertain whether the relationship between adversity and mental health was moderated by socio-economic position (SEP).

## Methods

### Participants

We use data from the COVID-19 Social Study; a large panel study of the psychological and social experiences of over 70,000 adults (aged 18+) in the UK during the COVID-19 pandemic. The study commenced on 21 March 2020 and involves online weekly data collection from participants for the duration of the pandemic in the UK. The study is not random but does contain a well-stratified sample with good representation across all socio-demographic groups (see Supplementary Material for further information on recruitment). The study was approved by the UCL Research Ethics Committee [12467/005] and all participants gave informed consent. Full details of the study, recruitment, retention, protocol and user guide are available at www.covidsocialstudy.org

As questions asked about experiences of adversity in the last week, we focused on data 1^st^ April 2020 (one week after lockdown commenced) to 12^th^ May 2020, limiting our analysis to participants who were interviewed on two or more occasions during this period (n = 48,723, observations = 208,057). We used complete case data, and excluded participants with complete data in fewer than two interviews (n = 6,814; 14.0% of eligible participants). This provided a final analytical sample of 41,909 participants (178,430 observations).

## Measures

### Depression

Depression during the past week was measured using the Patient Health Questionnaire (PHQ-9); a standard 9-item instrument for diagnosing depression in primary care, with 4-point responses ranging from “not at all” to “nearly every day” (range 0-27; higher scores indicate more depressive symptoms) ^18 19^.

### Anxiety

Anxiety during the past week was measured using the Generalised Anxiety Disorder assessment (GAD-7); a well-validated 7-item tool used to screen and diagnose generalised anxiety disorder in clinical practice and research, with 4-point responses ranging from “not at all” to “nearly every day” (range 0-21; higher scores indicate more symptoms of anxiety) ^20^.

### Adversities

We study six categories of adversity, each measured weekly (see Table 1). We constructed weekly total adversity worries and total adversity experiences measures by summing the number of adversities present in a given week (range 0-6). We considered worries to be one-off events and counted them only in the weeks they were reported. For adversities that are likely to be continuing (i.e. once experienced in one week, their effects would likely last into future weeks), we counted them on subsequent waves after they had first occurred. This applied to experiencing suspected/diagnosed COVID-19, loss of paid work, major cut in household income, and abuse victimisation.

**Table 1:** Questions on adversities

| Type of adversity | Adversity worries | Adversity experiences |
| --- | --- | --- |
| COVID-19 illness | Worried about catching COVID-19 | Currently have or previously had suspected or diagnosed COVID-19 |
| Financial difficulty | Worried about finances | Experienced a major cut in household income |
| Loss of paid work | Worried about losing your | Lost one's job or been unable to do |

|  | job/unemployment | paid work |
| --- | --- | --- |
| **Difficulties acquiring medication** | Worried about getting food | Unable to access sufficient food |
| **Difficulties accessing food** | Worried about getting medication | Unable to access required medication |
| **Threats to personal safety** | Worried about personal safety/security | Experienced being physically harmed or hurt by somebody else or being bullied, controlled, intimidate or psychologically hurt by someone else |

### Socio-Economic Position

We measured SEP using five variables collected at baseline interview: annual household income (<£16,000, £16,000 - £30,000, £30,000-£60,000, £60,000-£90,000, £90,000+), highest qualification (GCSE or lower, A-Levels or vocational training, undergraduate degree, postgraduate degree), employment status (employed, inactive, and unemployed), housing tenure (own outright, own with mortgage, rent/live rent free), and household overcrowding (binary: >1 persons per room). From these variables, we constructed a *Low SEP* index measure by counting indications of low SEP (income <£16k, educational qualifications of GCSE or lower, unemployed, living in rented or rent free accommodation, and living in over-crowded accommodation), collapsing into 0, 1, and 2+ indications of low SEP to attain adequate sample sizes for each category.

### Analysis

We used fixed-effects regression, which differs from other regression techniques as it explores within-person variation with individuals serving as their own reference point, compared with themselves over time. So all time-invariant (stable) covariates, are accounted for, even if unobserved ^21^. This approach is advantageous as individual stable characteristics such as socio-economic status, genetics, personality, history of mental illness and threshold for worries could confound associations between adversities and mental health. But as individuals are compared with themselves, such bias cannot affect results. Additionally, having experiences and worries varies over time, as does mental health, and both can be affected by time-varying confounders. Fixed-effects regression allows us to analyse these time-varying associations. So in this analysis we were able to assess the relationship between changes in both experiences and worries about adversity across six weeks of data collection, with changes in mental health.

In Model 1, we regressed each mental health measure on the total number of adversity experiences and total number of adversity worries, both (a) separately and (b) jointly, using the fixed effects estimator to account for time-invariant heterogeneity across participants. In Model 2, we regressed each measure of mental health on adversity experiences and adversity worries separately for each category of adversity in turn. In Model 3, we repeated Model 1a including interactions between adversity measures and the low SEP index, in order to estimate differences in associations by SEP. We adjusted for day of week (categorical) and days since lockdown commenced (continuous) in each regression, and we standardised GAD-7 and PHQ-9 scores to aid comparison across the two measures. Other time-constant confounders were automatically adjusted for due to the analytical approach. To account for the non-random nature of the sample, all data were weighted to the proportions of gender, age, ethnicity, education and country of living obtained from the Office for National Statistics ^22^. All graphs show standardised coefficients (predicted change in standardised Likert scores). Analyses were carried out in Stata version 16.0 (Statacorps, Texas) and RStudio version 3.6.3.

## Results

Table 2 provides detail on the demographic composition of our sample. Descriptive statistics for our exposures and outcomes are shown in Table S2. There was within-variation in each of the measures, suggesting fixed effects was a valid approach. Table S1 in the supplementary material displays descriptive statistics for PHQ-9 and GAD-7 scores by SEP group. Our sample showed clear social gradients in anxiety and depression symptoms.

**Table 2:** Sample demographics (n [%]), weighted and unweighted figures.

|  | Variable | Unweighted Data | Weighted Data |
| --- | --- | --- | --- |
| Age (grouped) | 18-34 | 6,336 (15.12%) | 6,947.33 (18.56%) |
|  | 35-49 | 12,893 (30.76%) | 9,630.17 (25.73%) |
|  | 50-64 | 14,060 (33.55%) | 11,734.81 (31.35%) |
|  | 65+ | 8,620 (20.57%) | 9,121.10 (24.37%) |
| Country of residence | England | 33,324 (79.52%) | 31,178.53 (83.29%) |
|  | Scotland | 2,835 (6.76%) | 3,179.33 (8.49%) |
|  | Wales | 5,303 (12.65%) | 2,246.93 (6%) |
|  | Northern Ireland | 447 (1.07%) | 828.63 (2.21%) |
| Ethnicity | White | 40,069 (95.61%) | 33,913.52 (90.6%) |
|  | BAME | 1,840 (4.39%) | 3,519.89 (9.4%) |
| Gender | Male | 10,711 (25.56%) | 18,528.02 (49.5%) |
|  | Female | 31,198 (74.44%) | 18,905.39 (50.5%) |
| Household income | £90k+ | 4,559 (10.88%) | 2,951.29 (7.88%) |
|  | £60k - £90k | 6,557 (15.65%) | 4,514.45 (12.06%) |
|  | £30k - £60k | 14,739 (35.17%) | 12,103.26 (32.33%) |
|  | £16k - £30k | 10,034 (23.94%) | 10,350.80 (27.65%) |
|  | <£16k | 6,020 (14.36%) | 7,513.60 (20.07%) |
| Marital status | Single | 6,579 (15.7%) | 6,983.13 (18.65%) |
|  | Divorced/Widowed | 5,691 (13.58%) | 4,974.23 (13.29%) |
|  | In relationship but living apart | 2,573 (6.14%) | 2,577.85 (6.89%) |
|  | Cohabiting with partner | 27,066 (64.58%) | 22,898.20 (61.17%) |
| Overcrowded accommodation | Not Overcrowded | 40,233 (96%) | 35,325.10 (94.37%) |
|  | Overcrowded | 1,676 (4%) | 2,108.31 (5.63%) |
| Highest qualification | Postgraduate | 11,893 (28.38%) | 5,591.47 (14.94%) |
|  | Undergraduate | 17,314 (41.31%) | 8,109.11 (21.66%) |
|  | A-Level or Vocational | 7,196 (17.17%) | 12,247.32 (32.72%) |
|  | GCSE or Lower | 5,506 (13.14%) | 11,485.50 (30.68%) |
| Employment status | Employed | 28,060 (66.95%) | 22,644.11 (60.49%) |
|  | Inactive | 13,402 (31.98%) | 14,226.83 (38.01%) |
|  | Unemployed | 447 (1.07%) | 562.47 (1.5%) |
| Household tenure | Own Outright | 14,985 (35.76%) | 12,954.54 (34.61%) |
|  | Own Mortgage | 15,926 (38%) | 11,969.65 (31.98%) |
|  | Rent | 10,998 (26.24%) | 12,509.22 (33.42%) |

Both the total adversities and total worries indices were associated with increases in GAD-7 and PHQ-9 scores (**Error! Reference source not found**.). The inclusion of worries in the same model as experiences slightly reduced the effect size of experiences, although the inclusion of experiences in the model had little effect on the effect size of worries. Effect sizes for number of experienced adversities were similar across depression and anxiety measures, but adversity worries were more highly related to anxiety symptoms than depression symptoms.

Worries about all types of adversities showed associations with higher levels of GAD-7 and PHQ-9 scores (Figure 2). Experiences of adversities relating to accessing food, accessing medication, and personal safety were also associated with higher levels of GAD-7 and PHQ-9 scores. However, experience of adversities relating to employment and finance were not associated with changes in mental health, and experience of COVID-19 symptoms was only related to higher depression scores. Experience of harm was more strongly related to mental health than worry about personal safety.

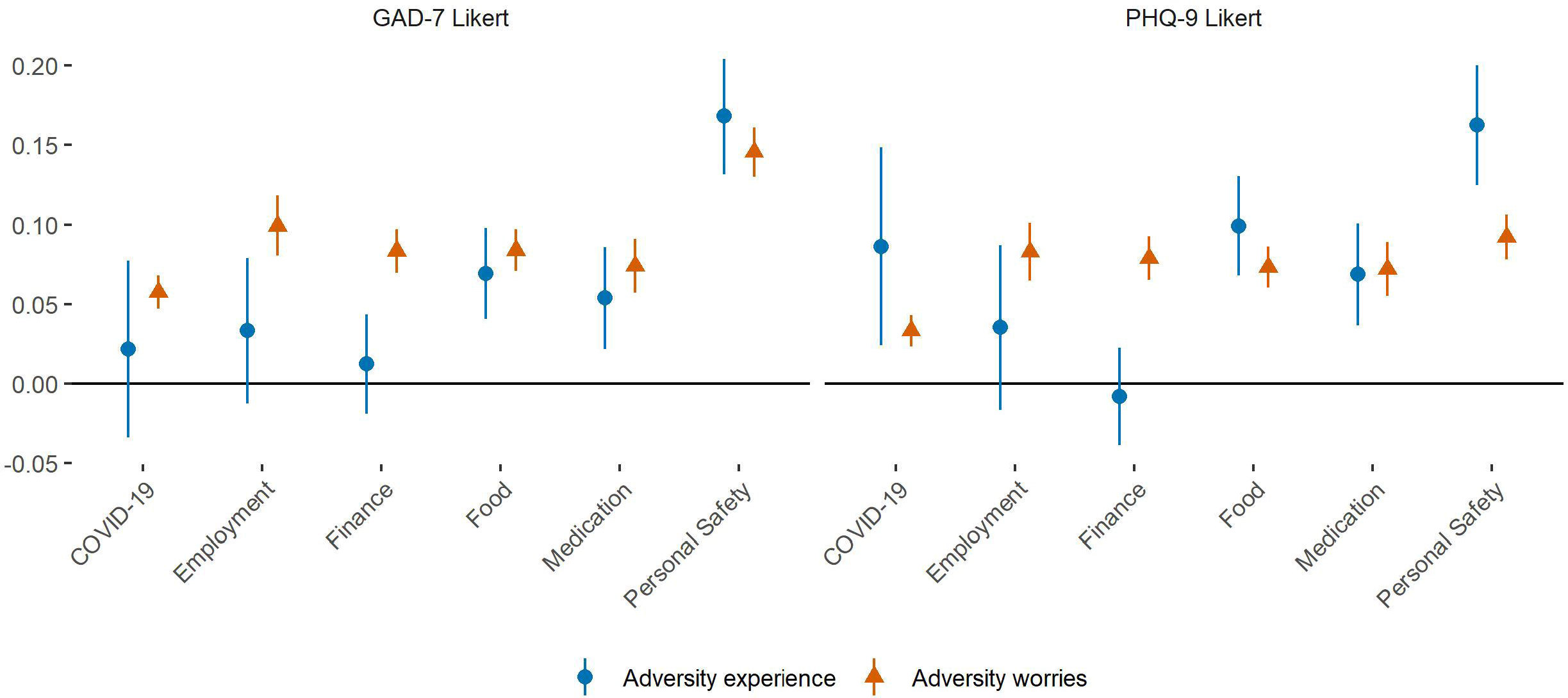

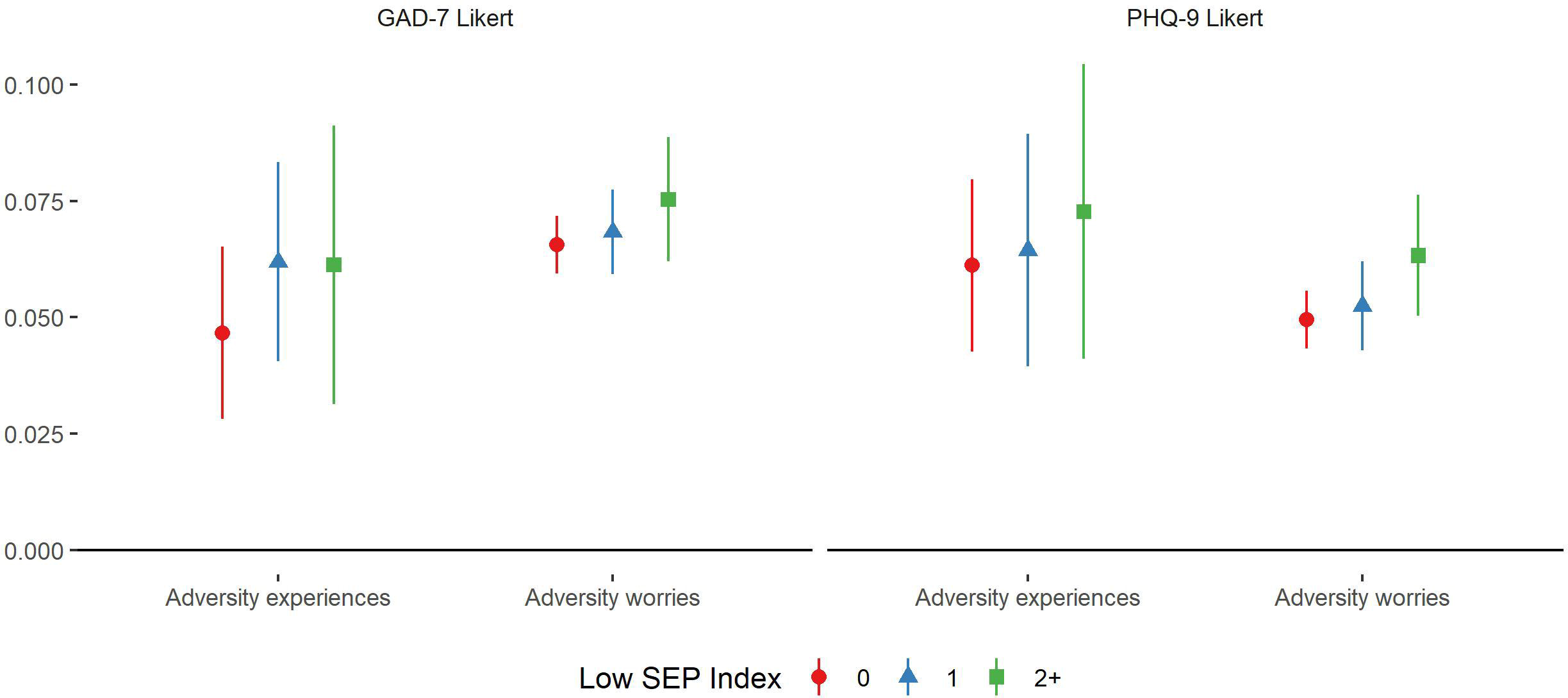

There was some evidence of a social gradient in the association between adversity experiences and adversity worries and mental health outcomes, with stronger associations found in more disadvantaged groups (Figure 3). However, individual estimates showed substantial variability, especially for experiences.

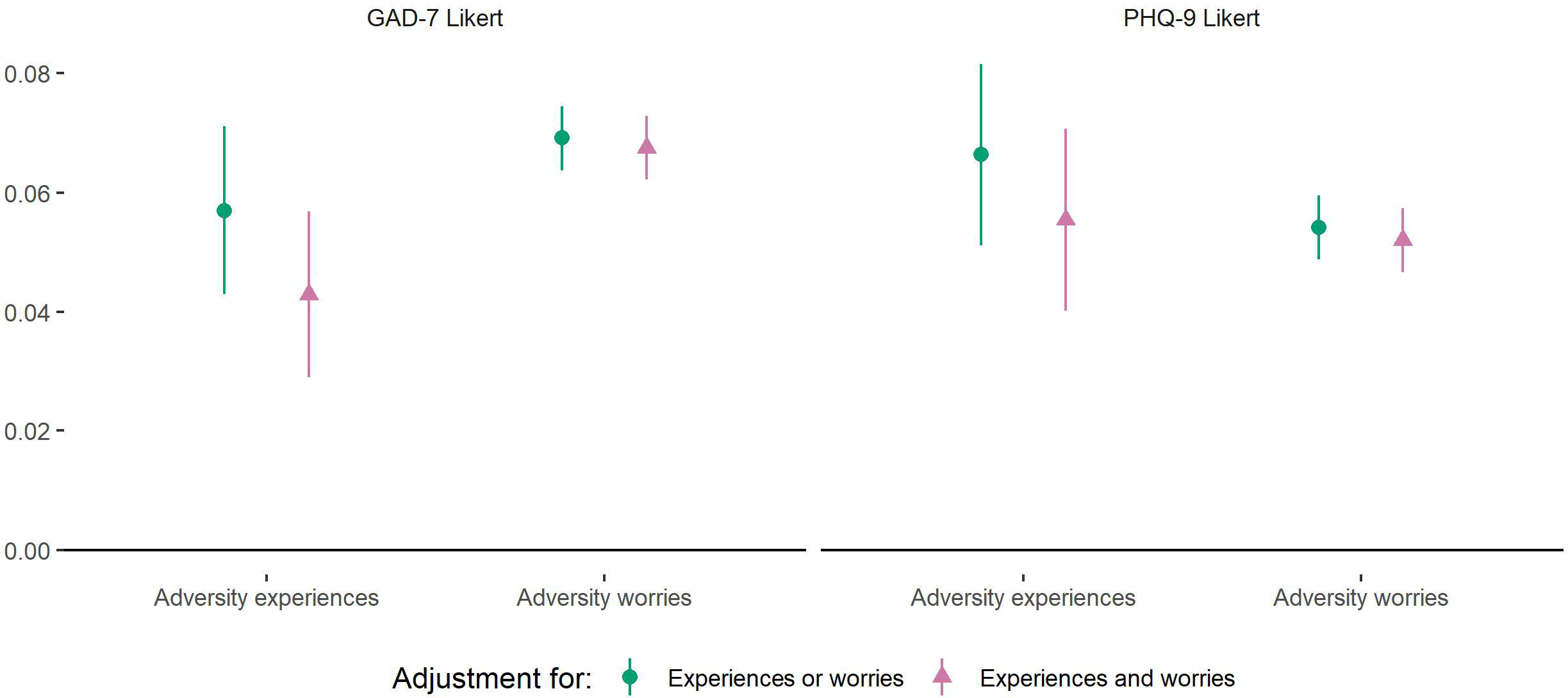

### Sensitivity Analyses

We carried out several sensitivity analyses to test the robustness of our results. When re-estimating Models 1 and 2 using inability to pay bills, rather than major cut in household income, as our measure of experienced financial adversity, the point estimates were more highly related to depression and anxiety symptoms (Figure S1 & S2). When repeating Models 1-3 using only reports of “significant stress” as opposed to minor stress to define the worries variables, effect sizes were predictably larger (Figures S3 and S4), and there was evidence of a reverse social gradient for the relationship between adversity worries and anxiety symptoms, with largest associations found among the least disadvantaged groups (Figure S5). To assess whether our findings were just an artefact of our categorisation of SEP, we re-estimated Model 3 using quintiles of a continuous measure of SEP derived from a confirmatory factor analysis (CFA) of the five SEP indicator variables (see Supplementary Material methods). There was still some weak indication of a social gradient in the association between adversity experiences and mental health, and some indication of a reverse social gradient in adversity worries and anxiety symptoms, with smaller associations found in more disadvantaged groups (Figure S6 and Table S3).

## Discussion

In this study, we explored the relationship between worries and experience of adversities and mental health during the first few weeks of the lockdown due to COVID-19 in the UK. Cumulative number of worries and experience of adversities were both related to higher levels of anxiety and depression. Number of worries were associated more strongly with anxiety than depression, but number of experiences were equally related to anxiety and depression. When considering specific types of adversities, there was greater variability in the relationship between experiences and mental health than worries and mental health. Worries were more strongly related to mental health than experiences for employment and finances, but less for personal safety and catching COVID-19. Individuals of lower SEP were more negatively affected psychologically by adverse experiences, but the relationship between worries, SEP and mental health was unclear.

Our findings that number of worries were more closely related to anxiety than to depression echoes previous research 23. Indeed, worrying is an integral component of many kinds of anxiety disorders 24, with substantial worrying or “rumination” associated with poor mental health ^25^. The finding that number of worries about adversities and number of experiences of adversity were equally related to anxiety echoes previous work highlighting how the impact worries about events can be equal to or even greater than experiences of events ^14^. The results on worries may indicate a bidirectional process between experiencing worries during COVID-19 and becoming more anxious. However, for depression there is less evidence of a bidirectional relationship in previous literature. Instead, reactivity to worries has been found to be a vulnerability factor for depression, but depression has not been found to predict higher negative reactivity ^10,26^.

In relation to experience of adversities, the fact that cumulative experiences was associated with poorer mental health but only certain specific experiences showed the same association suggests that it is the toll of cumulating events that is particularly challenging, perhaps as individual capabilities to manage challenging situations become exhausted ^11^. However, lack of immediate response to an adversity does not necessarily imply that mental health is not affected, as for certain worries, adverse consequences for mental health may take time to arise. For instance there is a reported delayed response time in mental health responses to unemployment ^27^, with rejections during job searches or cuts in income starting to impact on living standards appearing to be bigger triggers than the loss of work itself ^28^. In line with this, it is notable that we found higher associations for the relationship between inability to pay bills and mental health than loss of income and mental health. Indeed, while loss of income could occur across the wealth spectrum, inability to pay bills is likely concentrated at those with lower levels of household income, so could be regarded as a more significant experience. Financial adversities may also have been anticipated, which may have decreased mental health in the lead up to the event, leading to a floor effect by the time the event occurred ^27^. But it is also possible that the fear of potential adversity, in particular given the low levels of control experienced in worrying, is psychologically more demanding than the adjustment after an adverse event has occurred ^29^. The exception to this theory on psychological demand is experiences of adversities relating to personal safety. These were much more strongly linked with mental health than worries about personal safety, and had the strongest link with mental health out of all adversities assessed, which echoes research on the strong negative mental health impact of domestic abuse and violence ^30^.

In relation to catching COVID-19, there was a relationship between worries about catching the virus and anxiety, but there was much greater variability in the relationship between actually catching the virus and mental health. It is possible that there was selection bias in the study, with only individuals who caught and recovered from COVID-19 continuing to take part. But it is also possible that in terms of anxiety, the experience of the virus was less bad than some people had been fearing, leading to relief that individuals had not experienced serious health consequences. Nevertheless, although the confidence intervals were wide, there was still evidence to suggest that catching COVID-19 was associated with increases in depression. This is interesting given evidence suggesting that COVID-19 leads to the release of pro-inflammatory cytokines associated with depressive disorders ^31^, and remains to be explored further in future research.

There was some slight evidence of a stronger relationship between adverse experiences and both anxiety and depression amongst people of low SEP. This echoes previous research suggesting that higher SEP can be a buffer against the effects of adversity, with individuals of lower SEP more vulnerable especially to economic shocks ^17^. But it is also of note that there was some evidence of a reverse social gradient for adversity worries (especially for more significant worries), with individuals of higher SEP more affected. This could suggest that people who usually face fewer adversities in day to day life, the experience of new worries relating to adversity may have more profound effects ^32^. Or it could reflect the already higher levels of anxiety and depression found amongst individuals of lower SEP, suggesting a ceiling effect in reactivity to stressful situations ^3,15^.

This study had a number of strengths including its large, well-stratified sample, which was weighted to population proportions for core socio-demographic characteristics. Further, the study collected data covering the entire period from the start of lockdown in the UK on a weekly basis, providing an extremely rich dataset with longitudinal data. Our statistical approach (fixed effects regression) also allowed the comparison of individuals against themselves (within rather than between-subjects comparisons), so changes over time in the experience of worries and mental health were relative to each individual. As such, our measurement of worries was relative to each individual’s own perspectives, circumstances and coping threshold, allowing us to assess changes in an individual’s perception of their worries over time. Although, it should be noted that there were much wider confidence intervals measurements of association between experiences and mental health compared to worries and mental health, suggesting that people’s responses to experiences are much more variable, presumably due to differences in coping styles and wider circumstances. However, the study had several limitations. Our sampling was not random, so although we deliberately sampled from groups such as individuals of low SEP and individuals with existing mental illness, it is possible that more extreme experiences were not adequately captured in the study. It is also possible that individuals experiencing particularly extreme situations during the lockdown withdrew from the study. While our statistical method means their data is still included, we would lack longitudinal follow-up on their changing experiences. We also focused on just six types of adversities, including those relating to health, safety, finances and basic needs. However, many other types of adversity were not included in the study, including those relating to interpersonal relationships, displacement, and bereavement. Finally, our study only followed individuals up for a few weeks looking at the immediate associations with mental health. As such, it remains for future studies to assess how experience of adversities during the COVID-19 pandemic relates to long-term mental health consequences.

Overall, the finding that mental health was associated both with experiences and worries about adversities suggests that interventions are required that both seek to prevent adverse events (such as loss of jobs) but also that reassure individuals and support adaptive coping strategies. This appears to be particularly important for managing anxiety, where provision of online cognitive-behavioural training may help support individuals in the management of uncertainty. These results suggest that measures over the first few weeks of lockdown in the UK have been insufficient at reassuring people given we are still seeing clear associations with poor mental health both for cumulative worries and also for a range of specific worries relating to finance, access to essentials, personal safety and COVID-19. Given the challenges in providing mental health support to individuals during the lockdown, these findings highlight the importance of developing online and remote interventions that could provide such support, both as COVID-19 continues and in preparation for future pandemics.

## Data Availability

Data used in this study will be made publicly available once the pandemic is over.

## Acknowledgements

The researchers are grateful for the support of a number of organisations with their recruitment efforts including: the UKRI Mental Health Networks, Find Out Now, UCL BioResource, HealthWise Wales, SEO Works, FieldworkHub, and Optimal Workshop.

## Author contribution

LW, AS and DF conceived and designed the study. LW analysed the data and DF wrote the first draft. All authors provided critical revisions. All authors read and approved the submitted manuscript.

## Data availability

Data used in this study will be made publicly available once the pandemic is over.

